# Estimating levels and trends in labour induction worldwide: a systematic review and modelling analysis

**DOI:** 10.64898/2026.05.20.26353632

**Authors:** Samia Aziz, Yanan Hu, Saima Sultana, Neasha Jayakody, Breanna Teo, Elizabeth Korevaar, Amalia Karahalios, Fiona Bruinsma, Caroline SE Homer, Joshua P Vogel

## Abstract

**Introduction:** Induction of labour is a widely used obstetric intervention, yet its use varies markedly, with underuse in some settings and increasing elective use in others. However, the global prevalence and trends worldwide is unknown. We aimed to synthesise national and subnational data to estimate the prevalence of labour induction internationally and assess trends over time.

**Methods:** We sought data from 194 countries through a structured search of national databases and relevant websites. For countries lacking adequate national data, we conducted a systematic review of published studies. Eligible data were pooled to estimate the prevalence of labour induction for 2019, and to examine temporal trends from 2010 to 2022. We used mixed-effects negative binomial regression models with missing data handled using multiple imputation by chained equations.

**Results:** Data were obtained for 62 countries, including national-level data from 19 countries and 176 studies from 43 countries. Overall, 40 countries contributed to the 2019 estimate and 43 to the trend analysis. Most countries with data were high-income (N=37, 86.0%) and in Europe (N=29, 67.4%); there were no eligible data for sub-Saharan Africa. The estimated rate of labour induction for 2019 was 23·7% (95% confidence interval (CI): 19·3% to 29·2%). Induction had an estimated annual increase of 4% between 2010 and 2022 (incidence rate ratio 1·04, 95% CI 1·02 to 1·06).

**Conclusion:** This study provides the first international estimates of labour induction, revealing high and rising rates globally. These trends likely reflect expanded clinical indications and improved access, but also signal potential overuse in resource-rich contexts. Our findings highlight a critical ‘data gap’ in LMICs, particularly in Sub-Saharan Africa. Strengthening national perinatal data systems, especially in these settings, is essential for monitoring and guiding appropriate use. Identifying the optimal induction rate should be a priority for future research and clinical practice.

## INTRODUCTION

Induction of labour is the process of initiating labour before it begins spontaneously.^1^ The practice originated in ancient Egypt, where natural remedies such as castor oil were used.^2^ Labour induction was used in the late 18^th^ century in the United Kingdom (UK) to manage a ‘contracted pelvis’, then considered a potentially fatal condition.^3^ In contemporary clinical practice, it is a common intervention that has benefits for woman and baby. Clinical indications include its use in post-term pregnancy to reduce the risk of perinatal mortality after 41 weeks’ gestation.^4^ It is also recommended for the management of prelabour rupture of membranes, fetal growth restriction, hypertensive disorders of pregnancy, and gestational diabetes.^1,5^ Induction is not without risk, and can contribute to shoulder dystocia, need for caesarean section, or uterine rupture (particularly if a woman’s uterus is scarred).^4^ Evidence from multicounty trials and large observational studies demonstrating benefits of induction of labour at term in low-risk women over expectant management^6-8^ has contributed to its increasing use in many countries.^9^ This rising use has raised concerns about the over-medicalisation of birth and increased health system costs.^10^ Whereas settings with low or minimal induction use may reflect missed opportunities to improve maternal and perinatal outcomes.

In some high-income countries, national data show approximately one in every four births take place following labour induction.^1^ In Australia, induction use increased from 26% in 1990 to 33% in 2023; similar increases have been observed in the UK and the United States of America (USA).^11-13^ Many countries do not have national perinatal data systems,^14^ or such systems may not routinely capture labour induction data. An analysis of a World Health Organization (WHO) multicountry survey conducted in 23 low- and middle-income countries (LMICs) found labour rates ranging from 1·4% in Niger to 6·8% in Algeria across seven African countries (2004–05), and from 2-5% in Cambodia to 35·5% in Sri Lanka amongst eight Asian countries (2007-08).^15,17^

Understanding patterns of induction use internationally is critical for global maternal and newborn health, particularly in the context of the Sustainable Development Goals and other global targets to reduce preventable maternal and perinatal mortality, including stillbirths.^16,17^ These data are essential to understanding global and regional patterns in induction use, monitoring adherence to evidence-based recommendations for induction, and aligning policy with clinical practice gaps. The aim of this study was to estimate the levels and trends in induction of labour globally, through a systematic review and pooled analysis of available data.

## METHODS

The study protocol was registered with PROSPERO (CRD42023490778) on 6 December 2023. The protocol was developed *a priori*, and the systematic review was conducted and reported in accordance with the PRISMA 2020 guidelines (Preferred Reporting Items for Systematic Reviews and Meta-Analyses)^18^ (Appendix Table Sla, Sib), with relevant adaptations appropriate for a prevalence review.

### Search strategy and selection criteria

We systematically searched for data on the induction of labour from 1 January 2010 onwards, for all 194 WHO Member States. Our preferred data source was national-level data from governments. We used a two-stage approach – stage 1 was a structured search for national-level data sources reporting induction of labour, and stage 2 was a systematic review of published studies with labour induction data, confined to countries where stage 1 national-level data were unavailable or insufficient.

In stage 1, we directly searched the websites of Ministries of Health and National Statistics Offices of all countries, seeking publicly available reports or data on induction of labour. We were aided by the United Nations portal containing links to all National Statistics Offices websites.^19^ In addition, we conducted targeted web searches with predefined search terms (e.g., *perinatal report, maternal health report, birth registry, induction of labour)* to identify global and regional maternal and perinatal health reports containing population-level data for individual countries, such as the European Perinatal Health Report.^20^ If any uncertainties arose regarding government data (e.g., data definitions, methodological details or missing years), we contacted the relevant country authorities to seek clarification and, where possible, obtained more detailed data. Stage 1 searches were conducted between 10 August 2023 and 31 January 2024. All reports were collated and data were extracted into a pretested Excel form. Extraction was performed by a first reviewer (SA), with a second reviewer (NJ, FB, YH) independently verifying extracted data by cross-checking against the source document. Data fields included country, data source, population, sample size, study period, gestational age, parity, induction of labour data (number of births/live births, number of inductions of labour, proportion/rate of induction of labour) and method/s of induction.

For stage 2, we identified the 175 countries where stage 1 national-level data were unavailable or where stage 1 data were insufficient. Insufficient data were defined as fewer than seven annual data points for the 13-year period from 2010 to 2022 (i.e. <50% coverage for this time period). We conducted a systematic review of published studies to identify population-based or facility-based studies reporting labour induction data in these countries. The following electronic databases were searched: MEDLINE, EMBASE and PsycINFO (via Ovid) and CINAHL and Scopus (via EBSCOhost). A search strategy was developed (Appendix pp 9-12) with consultation with a librarian, and the last search was conducted on 15 June 2024. The search was restricted to studies published from 2010 onwards, with no language restrictions. Eligible studies for stage 2 were those reporting induction use in a population of all births or live births, among mixed-risk, low-risk, or otherwise unselected-risk pregnancies. We did not include data from studies using selected populations of high-risk women only (Appendix p 12), given induction of labour would be artificially high in these subgroups. We included only those studies with at least 5,000 births to minimize small-study effects on prevalence estimation. We considered observational studies as eligible, and excluded randomised controlled trials, systematic reviews, case-control studies or case reports.

All records recovered from these searches were imported into Covidence and deduplicated.^21^ Two reviewers (SA and one of SS, YH, NJ or BT) independently screened titles and abstracts for all unique records to identify potentially eligible studies. Full texts of these were obtained and screened independently by two reviewers, using the eligibility criteria. Disagreements were resolved via discussion or through consultation with a third reviewer. We further scrutinized these studies, excluding those published after 2010 where the reported data were collected before 2010. If the study had a multi-year period, we included only those where at least part of the study period overlapped with 2010 or later. Data were extracted using a pre-tested Excel form by one author (SA) and independently checked by a second author (YH, SS or NJ). Any discrepancies identified during the verification process were resolved through discussion. The data extracted for stage 2 were similar to the stage 1 data fields, including year-specific data (where available) as well as cumulative data for the full study period (i.e., total number of inductions of labour and total number of births across all years).

In stage 2, we identified multiple studies reporting data for the same country and year. To minimize the risk of bias due to duplicative data sources, we applied a predefined hierarchical selection process for all studies in a single country, to retain a single data source per country-year (details provided in Appendix p 13-14). We assembled a master dataset comprising all extracted data from stages 1 and 2. Countries were classified using the World Bank income group (2023)^22^ and WHO region.

### Data analysis

We estimated the overall rate of labour induction for 2019 and examined temporal trends from 2010 to 2022 using all available data. All analyses were performed using Stata version 18.^23^ The Guidelines for Accurate and Transparent Health Estimates Reporting (GATHER) statement was followed in developing the dataset, analysis and presentation of results (Appendix Table S2).^24^ The year 2019 was selected as it represented the most recent year with the highest number of countries reporting labour induction data. This included countries with induction data reported for 2019, as well as countries where 2019 was included within a multiple-year study period (i.e., total number of inductions and the total population across all years in the study period).

The 2019 estimate was derived using a mixed-effects negative binomial regression model with the annual number of inductions of labour as the outcome, the yearly number of births as the exposure, and a random intercept for country to account for between-country heterogeneity. For the primary (complete-case) analysis, only countries with data for 2019 were included. Next, we fitted the same analytic model as the primary analysis and handled missing data using multiple imputation by chained equations with 40 imputed datasets. For this imputation model, year-specific numbers of inductions of labour and births were imputed using negative binomial regression within a fully conditional specification framework. Fixed terms for country were also included. Studies were eligible for inclusion in the imputation model if at least one annual induction rate could be calculated, or if a total induction rate was available for a multi-year period. In the latter case, the annual number of inductions and births were assumed constant across the study period. We explored inclusion of additional covariates or auxiliary variables which could explain the missing values; however, we could not identify variables to meaningfully inform imputation.

To estimate the annual rate of change, we fitted mixed-effects negative binomial regression models, with a fixed effect for year as a continuous variable and random intercepts for country. Robust standard errors were used to account for autocorrelation and heteroscedasticity. First, we handled missing data using complete-case analysis with year included as a continuous variable (Model 1). Next, missing year-specific data were imputed using multiple imputation with chained equations. The imputation model used negative binomial regression, with fixed effects for country and year as a continuous covariate (Model 2).

### Sensitivity analyses

To assess if our results were robust to the method for handling missing data, we compared the results with a multiple imputation model using Poisson regression (Model 3). To evaluate whether trends varied across countries, we extended this model by allowing random slopes for year, thereby permitting country-specific temporal trends (Model 4). In the multiple imputation framework, Poisson regression was used in the imputation stage, followed by mixed-effects negative binomial regression, as negative binomial models in the imputation stage did not converge. Finally, to assess the robustness of the imputation approach, we estimated the random slopes model using complete-case data only (Model 5), including countries with observed data without imputation.

To visually examine our assumption that the temporal change in the number of inductions per year was linear, we fitted a mixed-effects negative binomial model with the annual number of inductions as the outcome, annual births as the exposure, year entered as a categorical fixed effect, and a random intercept for country.

## RESULTS

Stage 1 searches identified national data for 26 countries - 22 in the WHO European region, two in the Americas, and two from the Western Pacific Region. These were all high-income countries. Of these, 19 countries were eligible for inclusion for analysis (Appendix Table S3), contributing 203 datapoints (Figure 1). The systematic review (stage 2) identified 176 eligible studies, of which 80 were included for analysis after applying the predefined selection approach. These 80 studies contained data for 43 countries. We combined data from stages 1 and 2 for a final dataset of 62 countries: 39 high-income countries, nine upper-middle-income countries, 11 LMICs, and three low-income countries. Of these countries, 29 were from the European region, nine from the Americas, eight from the Western Pacific, five from the Eastern Mediterranean region and five from South-East Asia, and six from sub-Saharan African countries.

**Figure 1.**
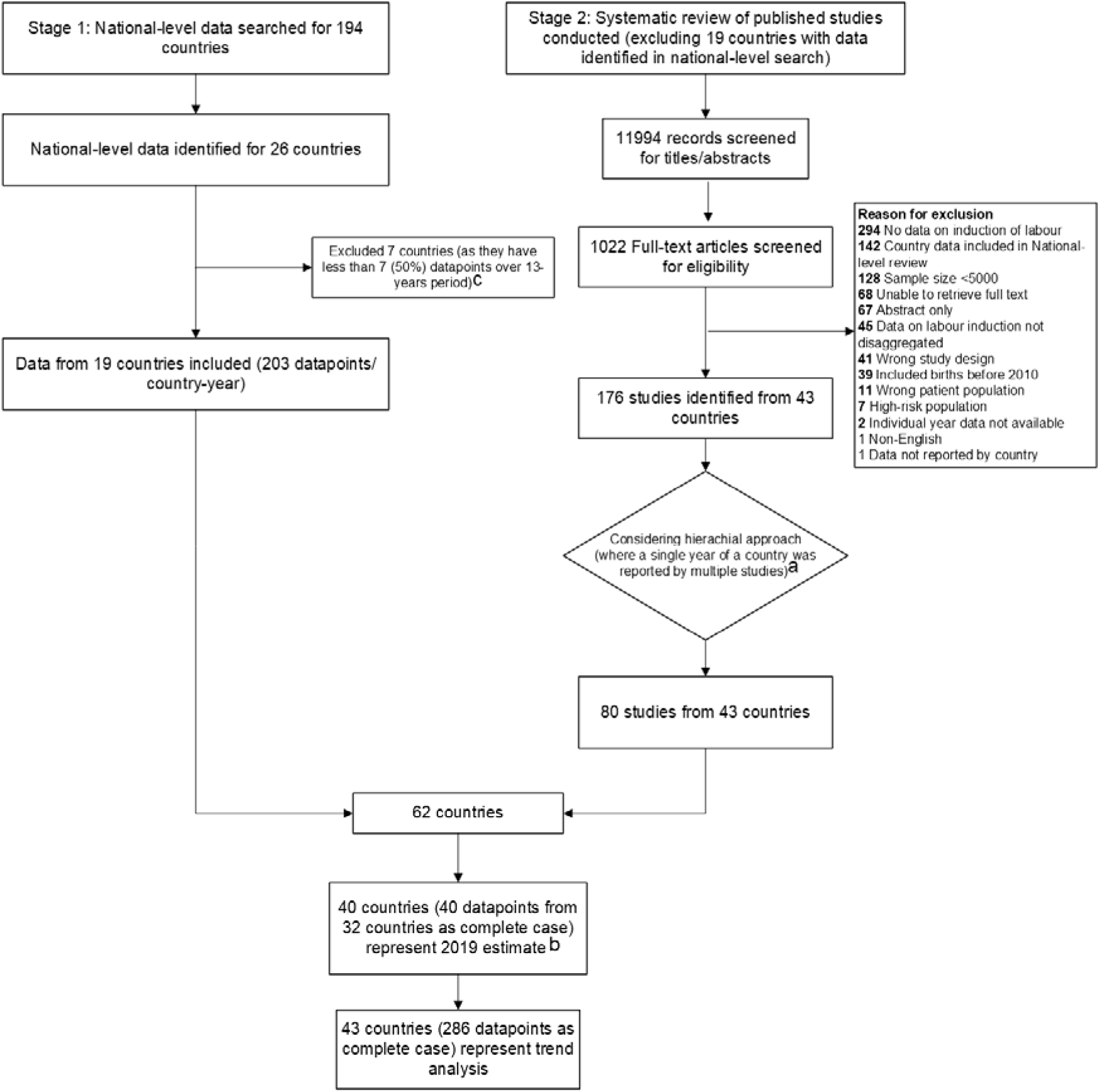
Identification of eligible data on induction of labour from national databases and a systematic review. ^a^When multiple studies reported data for the same country and year, a hierarchical selection approach was applied for inclusion in analysis dataset (box 1) ^b^ 2019 estimate for 32 countries (40 datapoints as complete-case) and an additional 8 countries included after multiple imputation (total 40 countries) ^c^ Seven countries were excluded from stage 1 due to insufficient data points. However, at Stage 2, additional individual-year and period data covering a longer time span were identified for these countries from studies that used national-level data. Therefore, the Stage 2 datapoints were included in the analysis for these seven countries.

From this final dataset (62 countries), 40 countries contributed data to the 2019 pooled estimate and 43 countries (37 high-income, four upper-middle-income and two LMICs) to the trend analysis (Appendix Table S4). Notably, no eligible data were available from countries in sub-Saharan Africa for inclusion in the analysis. Figure 2 illustrates the geographic distribution of countries included in the 2019 pooled estimate of induction of labour (Panel A) and the temporal trend analysis from 2010–2022 (Panel B).

**Figure 2.**
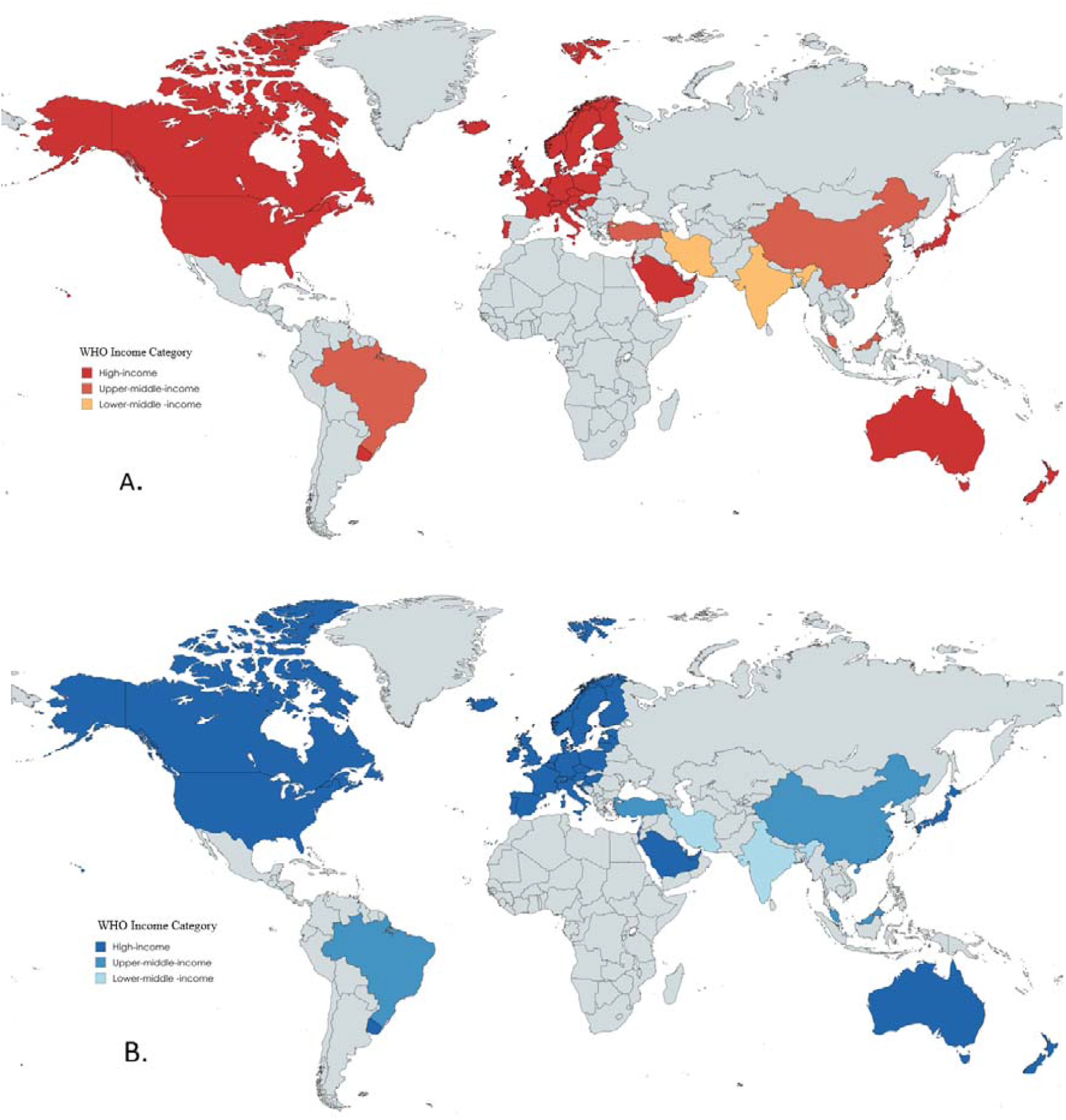
Countries included in the estimation of (A) induction of labour rates for 2019 and (B) temporal trends in induction of labour, 2010-2023. Panel A includes 40 countries which contributed data to estimate labour induction in 2019. Panel B shows the 43 countries that contributed data to estimate labour induction for 2010-2022. *\*Three additional countries included in Panel B are Belgium, Cyprus, and Spain*.

### Estimate of the prevalence of labour induction for 2019

A total of 32 countries (40 datapoints) contributed to the 2019 pooled estimate in the complete-case analysis. Induction of labour rates varied across these countries, ranging from 9·1% in the Czech Republic^25^ to 34·7% in Australia.^26^ Table 1 presents the estimated rate of labour induction for 2019 using both complete-case analysis and multiple imputation to handle the missing data. The complete-case analysis resulted in a pooled induction rate for 2019 of 23.3% (95% CI 20.1% to 26.9%). We imputed 2019 induction rates for an additional eight countries (40 countries total), finding a similar pooled estimate (23·7%; 95% CI 19·3% to 29·2%).

**Table 1.**
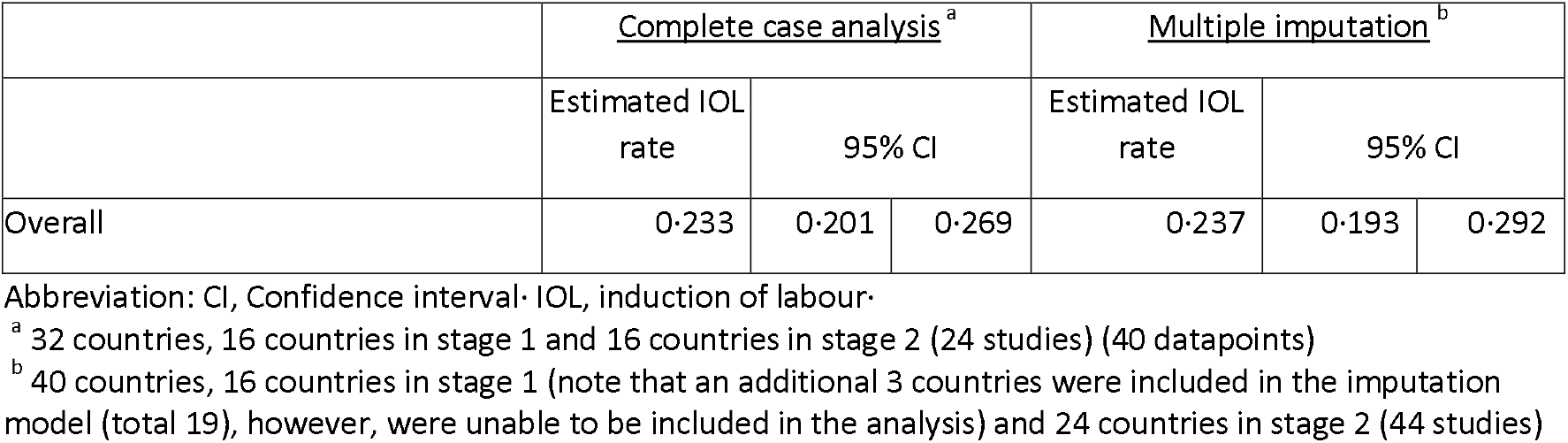
Estimated induction of labour use in 2019.

|  | <u>Complete case analysis<sup>a</sup></u> |  |  | <u>Multiple imputation<sup>b</sup></u> |  |  |
| --- | --- | --- | --- | --- | --- | --- |
|  | Estimated IOL rate | 95% CI |  | Estimated IOL rate | 95% CI |  |
| Overall | 0.233 | 0.201 | 0.269 | 0.237 | 0.193 | 0.292 |

Abbreviation: CI, Confidence interval; IOL, induction of labour.
<sup>a</sup> 32 countries, 16 countries in stage 1 and 16 countries in stage 2 (24 studies) (40 datapoints)
<sup>b</sup> 40 countries, 16 countries in stage 1 (note that an additional 3 countries were included in the imputation model (total 19), however, were unable to be included in the analysis) and 24 countries in stage 2 (44 studies)

### Trends of labour induction for 2010 to 2022

The complete case analysis included 43 countries (286 datapoints). The estimated labour induction rate increased annually (Incidence Rate Ratio (IRR) 1·04, 95% CI 1·02 to 1·05, Model 1) (Table 2). This estimate did not materially change when we used multiple imputation to handle the missing data (IRR 1·04, 95% CI 1·02 to 1·06, Model 2) (Table 2). Similar results were seen using a different method of multiple imputation, complete-case analysis, and a mixed-effect negative binomial model with random slopes (Models 3-5, Table 3). Although the overall trend was summarised as an average annual increase, annual estimates showed modest year-to-year variability around this underlying upward pattern (Figure 3).

**Table 2.** Estimated use of induction of labour over time.

|  | <u>Model 1<sup>a</sup></u> |  |  | <u>Model 2<sup>b</sup></u> |  |  |
| --- | --- | --- | --- | --- | --- | --- |
|  | Estimate | 95% CI |  | Estimate | 95% CI |  |
| Induction of labour rate in 2019 | 0.221 | 0.189 | 0.258 | 0.230 | 0.187 | 0.2829 |
| Incidence rate ratio per year | 1.035 | 1.024 | 1.047 | 1.036 | 1.016 | 1.057 |

Abbreviation: CI, Confidence interval.
<sup>a</sup> Model 1, Estimated from a mixed-effect negative binomial model with complete case analysis. Based on 43 countries, 19 in stage 1 and 24 in stage 2 (286 datapoints)
<sup>b</sup> Model 2, Multiple imputation using negative binomial regression, followed by mixed effect negative binomial model. Based on 43 countries, 19 in stage 1 and 24 in stage 2 (41 studies)

**Table 3.** Sensitivity analysis to estimate induction of labour over time.

|  | <u>Model 3<sup>a</sup></u> |  |  | <u>Model 4<sup>b</sup></u> |  |  | <u>Model 5<sup>c</sup></u> |  |  |
| --- | --- | --- | --- | --- | --- | --- | --- | --- | --- |
|  | Estimate | 95% CI |  | Estimate | 95% CI |  | Estimate | 95% CI |  |
| Induction of labour rate in 2019 | 0.209 | 0.178 | 0.245 | 0.210 | 0.179 | 0.247 | 0.216 | 0.185 | 0.252 |
| Incidence rate ratio per year | 1.032 | 1.028 | 1.036 | 1.034 | 1.030 | 1.038 | 1.029 | 1.018 | 1.041 |

Abbreviation: CI, Confidence interval.
<sup>a</sup> Model 3, Multiple imputation using Poisson regression, followed by mixed effect negative binomial model. Based on 43 countries, 19 countries in stage 1 and 24 countries from 41 studies in stage 2
<sup>b</sup> Model 4, Multiple imputation using Poisson regression, followed by mixed effect negative binomial model with random slopes. Based on 43 countries, 19 countries in stage 1 and 24 countries from 41 studies in stage 2
<sup>c</sup> Model 5, Estimated from a mixed effect negative binomial model with random slopes with complete case analysis. Based on 43 countries, 19 countries in stage 1 and 24 countries from 41 studies in stage 2 (41 studies)

**Figure 3.**
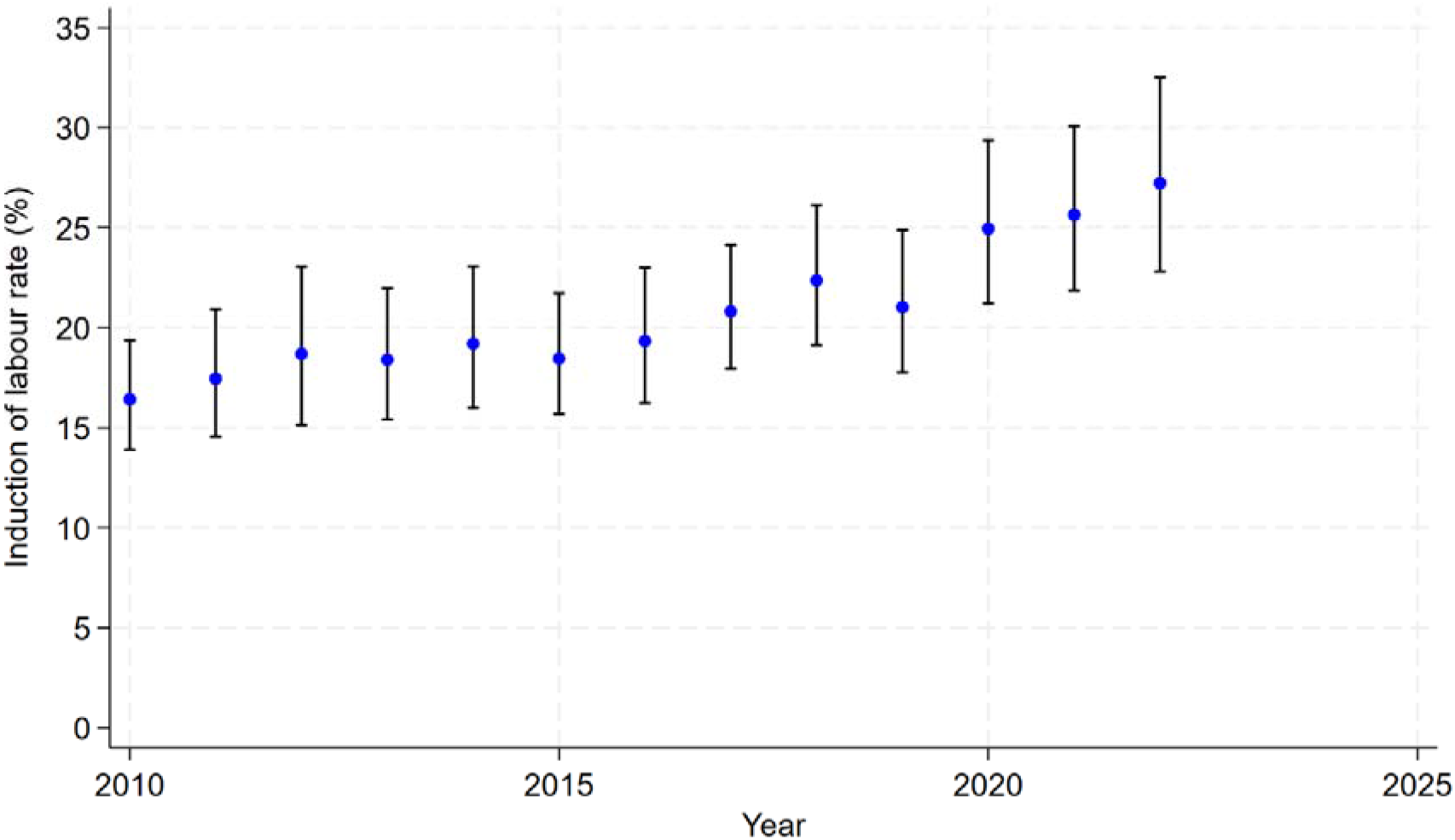
Annual induction of labour rates (%) (blue dots) and 95% confidence intervals (black bars) from a mixed-effect negative binomial regression model with annual number of inductions of labour as the outcome of interest, the population per year as the exposure, fixed effect for year (categorical), random intercepts for country, and robust standard errors.

## DISCUSSION

This study provides the first comprehensive synthesis of global data on labour induction, combining national-level and peer-reviewed data from 62 countries for a 10-year period. We found substantial data gaps on induction use, with limited data from LMICs, and no data from sub-Saharan African countries were eligible for inclusion in the analysis. Pooled estimates indicate that in 2019, approximately one in four births (23·7%, 95% CI 19·3% to 29·2%) occurred following labour induction. Trend data from 43 countries shows rising labour induction use, with an average annual increase of 4% (IRR 1·04, 95% CI 1·02 to 1·06).

Our findings highlight substantial differences in data availability and reporting across country income groups. High-income countries, particularly those in Europe, contributed the majority of available data, and consistently reported high and rising rates of induction of labour. In contrast, for most low-income countries we did not identify any national-level data. Where data were available, these were derived from subnational studies. The absence of high-quality, population-level data in many LMICs makes it challenging to characterize labour induction practices in these countries, and thus globally. This imbalance affects the representativeness of our pooled estimates, and highlights the persistent gaps in routine perinatal data systems in lower-income settings.^27,28^

Population-based household surveys like Demographic Health Surveys, Multiple Cluster Indicator Surveys, and Reproductive Health Surveys are used for perinatal and obstetric health data in many LMICs where civil registration vital statistics systems are not present or do not capture detailed birth data. However, these household surveys do not collect data on labour induction, despite its importance in clinical practice. Also, some national perinatal surveillance systems have been discontinued due to funding constraints, and the US Government defunded the Demographic Health Surveys program in 2025.^29^This represents a critical missed opportunity, particularly in settings with high perinatal mortality rates.^30^ In such contexts, appropriate and timely labour induction can be life-saving.^1,31^

The observed rise in labour induction over time is consistent with trends reported previously within individual high-income countries. For example, we have previously documented an annual increase in labour induction of 5·5% in Victoria, Australia’s second most populous state.^32^ Similarly, substantial rises have been found in the UK and USA.^13,33^ This raises questions about the appropriateness of current labour induction use, and whether potential overuse (or misuse) is present. Much of this rise is presumably driven by increased use of elective or routine induction at term.^34,35^ Although evidence on the benefits of elective induction remains mixed,^36^ particularly for low-risk women, the practice has expanded substantially in recent years. Induction of labour is shaped not only by clinical decision-making but also by women’s preferences, underscoring the importance of how information about induction is communicated and understood.^37^ The quality and consistency of counselling provided to women are likely to vary across settings, potentially influencing both demand for, and uptake of induction. This pattern warrants further investigation to better understand the magnitude of elective induction and to ensure that induction practices remain evidence-based, equitable, and appropriate.

In terms of strengths, this study represents the first comprehensive attempt to estimate the global rate of labour induction, synthesising national and subnational data using a rigorous, multi-stage search strategy encompassing 194 countries. By combining national-level data and published studies, it provides the strongest evidence to date on the global landscape of labour induction practices, and the extent of data gaps. While the pooled analysis contained data from only 43 countries, these countries collectively represent approximately 50% of the world’s population^38^ and 24% of the world’s total births in 2019 ^39^ Our statistical approach, including the use of mixed-effects negative binomial models, robust sensitivity analyses and multiple imputation, adds further methodological strength and ensures that the pooled estimate reflects the best available data. Although we initially aimed to estimate the rate of labour induction globally, data limitations meant that our pooled estimate was driven by high-income countries with relatively less contribution from LMIC data, limiting the generalisability of these findings. While multiple imputation improved the completeness of the trend analysis, the absence of suitable auxiliary variables specific to labour induction may have reduced precision of imputed values. From stage 2 studies, we selected a single study to represent each country-year (or mid-year) rather than averaging estimates across multiple studies. While this approach ensured consistency and prevented duplication of overlapping data sources, it may have excluded studies providing complementary subnational or regional coverage within the same country and year.

## CONCLUSION

The findings highlight the pressing need to strengthen national obstetric and perinatal surveillance and data systems, particularly in LMICs, and to standardise reporting of induction practices. Induction of labour is widely used and recommended internationally, and is life-saving in certain situations^1^ – it is thus important to ensure universal access. However, it is equally important to avoid overuse of labour induction, considering that it is not risk-free to woman and newborn, and may consume health resources that could be allocated elsewhere for better health gains. Future research should focus on how data systems can be improved to capture use of practices like induction, and ensuring that underrepresented countries are able to provide and evaluate evidence-based intrapartum care.

## AUTHOR CONTRIBUTIONS

The study was conceptualised by JPV and SA. The study protocol was written by SA with input from JPV, FB and CSEH. SA conducted data searches. SA, SS, YH, NJ, and BT screened, extracted and cross-checked data. AK and EK led the statistical analyses, which were reviewed by all named authors. SA drafted the first version of the manuscript. All authors reviewed and contributed substantially to data interpretation and manuscript revision, and approved the final version for submission.

## DECLARATION OF INTERESTS

We declare no competing interests.

## DATA AVAILABILITY STATEMENT

The dataset compiled for this analysis is based on publicly available national reports and published studies. A list of all stage 1 national data sources and the extraction sheet for studies included in stage 2 are provided in the Supplementary file.

## ACKNOWLEDGEMENTS

We acknowledge the contribution of Ms Lorena Romero (Alfred Hospital) in developing the search strategy. We also thank the Centers for Disease Control and Prevention (USA), the Directorate of Health, Iceland, and the Estonian Medical Birth Registry for providing additional information upon request.

## PATIENT AND PUBLIC INVOLVEMENT

Patients or the public were not involved in the design, or conduct, or reporting, or dissemination plans of our research.

## Notes

**Funding** Samia Aziz is supported by a Monash Graduate Research Scholarship through the School of Public Health and Preventive Medicine, Monash University. Prof Joshua P. Vogel is supported by an NHMRC Investigator Grant (GNT1194248), and Prof Caroline S.E. Homer is supported by an NHMRC Investigator Grant (APP1137745). Fiona Bruinsma is supported by NHMRC-funded ARPAN Centre of Research Excellence (GNT 2024658)(GNT2024658).

### Competing Interest Statement

The authors have declared no competing interest.

### Funding Statement

Samia Aziz is supported by a Monash Graduate Research Scholarship through the School of Public Health and Preventive Medicine, Monash University. Prof Joshua P. Vogel is supported by an NHMRC Investigator Grant (GNT1194248), and Prof Caroline S.E. Homer is supported by an NHMRC Investigator Grant (APP1137745). Fiona Bruinsma is supported by NHMRC-funded ARPAN Centre of Research Excellence (GNT 2024658)(GNT2024658).

