## Appendix for "Estimating levels and trends in labour induction worldwide: a systematic review and modelling analysis"

### 1. PRISMA 2020 Checklist

#### Table S1a PRISMA 2020 Checklist

| **Section and Topic** | **Item #** | **Checklist item** | **Location where item is reported** |
| --- | --- | --- | --- |
| **TITLE** | | |  |
| Title | 1 | Identify the report as a systematic review. | Page (P)1 |
| **ABSTRACT** | | |  |
| Abstract | 2 | See the PRISMA 2020 for Abstracts checklist. | Appendix file S1b |
| **INTRODUCTION** | | |  |
| Rationale | 3 | Describe the rationale for the review in the context of existing knowledge. | P 4 |
| Objectives | 4 | Provide an explicit statement of the objective(s) or question(s) the review addresses. | P 4 |
| **METHODS** | | |  |
| Eligibility criteria | 5 | Specify the inclusion and exclusion criteria for the review and how studies were grouped for the syntheses. | P 5,6 |
| Information sources | 6 | Specify all databases, registers, websites, organisations, reference lists and other sources searched or consulted to identify studies. Specify the date when each source was last searched or consulted. | P 5,6 |
| Search strategy | 7 | Present the full search strategies for all databases, registers and websites, including any filters and limits used. | Appendix, P 7-12 |
| Selection process | 8 | Specify the methods used to decide whether a study met the inclusion criteria of the review, including how many reviewers screened each record and each report retrieved, whether they worked independently, and if applicable, details of automation tools used in the process. | P 5,6 |
| Data collection process | 9 | Specify the methods used to collect data from reports, including how many reviewers collected data from each report, whether they worked independently, any processes for obtaining or confirming data from study investigators, and if applicable, details of automation tools used in the process. | P 5,6 |
| Data items | 10a | List and define all outcomes for which data were sought. Specify whether all results that were compatible with each outcome domain in each study were sought (e.g. for all measures, time points, analyses), and if not, the methods used to decide which results to collect. | P 5,6 |
|  | 10b | List and define all other variables for which data were sought (e.g. participant and intervention characteristics, funding sources). Describe any assumptions made about any missing or unclear information. | P 5 |
| Study risk of bias assessment | 11 | Specify the methods used to assess risk of bias in the included studies, including details of the tool(s) used, how many reviewers assessed each study and whether they worked independently, and if applicable, details of automation tools used in the process. | N/A |
| Effect measures | 12 | Specify for each outcome the effect measure(s) (e.g. risk ratio, mean difference) used in the synthesis or presentation of results. | P 6 |
| Synthesis methods | 13a | Describe the processes used to decide which studies were eligible for each synthesis (e.g. tabulating the study intervention characteristics and comparing against the planned groups for each synthesis (item #5)). | P 6,7  Appendix P 12-14 |
|  | 13b | Describe any methods required to prepare the data for presentation or synthesis, such as handling of missing summary statistics, or data conversions. | P 7 |
|  | 13c | Describe any methods used to tabulate or visually display results of individual studies and syntheses. | N/A |
|  | 13d | Describe any methods used to synthesize results and provide a rationale for the choice(s). If meta-analysis was performed, describe the model(s), method(s) to identify the presence and extent of statistical heterogeneity, and software package(s) used. | P 6,7 |
|  | 13e | Describe any methods used to explore possible causes of heterogeneity among study results (e.g. subgroup analysis, meta-regression). | P 6,7 |
|  | 13f | Describe any sensitivity analyses conducted to assess robustness of the synthesized results. | P 7 |
| Reporting bias assessment | 14 | Describe any methods used to assess risk of bias due to missing results in a synthesis (arising from reporting biases). | N/A |
| Certainty assessment | 15 | Describe any methods used to assess certainty (or confidence) in the body of evidence for an outcome. | N/A |
| **RESULTS** | | |  |
| Study selection | 16a | Describe the results of the search and selection process, from the number of records identified in the search to the number of studies included in the review, ideally using a flow diagram. | P 7,8 |
|  | 16b | Cite studies that might appear to meet the inclusion criteria, but which were excluded, and explain why they were excluded. | P 7, Supplementary file |
| Study characteristics | 17 | Cite each included study and present its characteristics. | Appendix, Table S3, Supplementary file |
| Risk of bias in studies | 18 | Present assessments of risk of bias for each included study. | N/A |
| Results of individual studies | 19 | For all outcomes, present, for each study: (a) summary statistics for each group (where appropriate) and (b) an effect estimate and its precision (e.g. confidence/credible interval), ideally using structured tables or plots. | Table 1-2 &Appendix Table S6  Figure 2-3 |
| Results of syntheses | 20a | For each synthesis, briefly summarise the characteristics and risk of bias among contributing studies. | P 7 |
|  | 20b | Present results of all statistical syntheses conducted. If meta-analysis was done, present for each the summary estimate and its precision (e.g. confidence/credible interval) and measures of statistical heterogeneity. If comparing groups, describe the direction of the effect. | P 7,8 |
|  | 20c | Present results of all investigations of possible causes of heterogeneity among study results. | P 7,8 |
|  | 20d | Present results of all sensitivity analyses conducted to assess the robustness of the synthesized results. | P 8 |
| Reporting biases | 21 | Present assessments of risk of bias due to missing results (arising from reporting biases) for each synthesis assessed. | N/A |
| Certainty of evidence | 22 | Present assessments of certainty (or confidence) in the body of evidence for each outcome assessed. | N/A |
| **DISCUSSION** | | |  |
| Discussion | 23a | Provide a general interpretation of the results in the context of other evidence. | P 8,9 |
|  | 23b | Discuss any limitations of the evidence included in the review. | P 9 |
|  | 23c | Discuss any limitations of the review processes used. | P 10 |
|  | 23d | Discuss implications of the results for practice, policy, and future research. | P 10 |
| **OTHER INFORMATION** | | |  |
| Registration and protocol | 24a | Provide registration information for the review, including register name and registration number, or state that the review was not registered. | P 4 |
|  | 24b | Indicate where the review protocol can be accessed, or state that a protocol was not prepared. | P 4 |
|  | 24c | Describe and explain any amendments to information provided at registration or in the protocol. | N/A |
| Support | 25 | Describe sources of financial or non-financial support for the review, and the role of the funders or sponsors in the review. | P 2 |
| Competing interests | 26 | Declare any competing interests of review authors. | P 10 |
| Availability of data, code and other materials | 27 | Report which of the following are publicly available and where they can be found: template data collection forms; data extracted from included studies; data used for all analyses; analytic code; any other materials used in the review. | P 10 |

*From:*  Page MJ, McKenzie JE, Bossuyt PM, Boutron I, Hoffmann TC, Mulrow CD, et al. The PRISMA 2020 statement: an updated guideline for reporting systematic reviews. BMJ 2021;372:n71. doi: 10.1136/bmj.n71. This work is licensed under CC BY 4.0. To view a copy of this license, visit [https://creativecommons.org/licenses/by/4.0/](https://creativecommons.org/licenses/by/4.0/ )

#### Table S1b. PRISMA 2020 for Abstracts Checklist

| **Section and Topic** | **Item #** | **Checklist item** | **Reported (Yes/No)** |
| --- | --- | --- | --- |
| **TITLE** | | |  |
| Title | 1 | Identify the report as a systematic review. | Yes |
| **BACKGROUND** | | |  |
| Objectives | 2 | Provide an explicit statement of the main objective(s) or question(s) the review addresses. | Yes |
| **METHODS** | | |  |
| Eligibility criteria | 3 | Specify the inclusion and exclusion criteria for the review. | Yes |
| Information sources | 4 | Specify the information sources (e.g. databases, registers) used to identify studies and the date when each was last searched. | Yes |
| Risk of bias | 5 | Specify the methods used to assess risk of bias in the included studies. | No |
| Synthesis of results | 6 | Specify the methods used to present and synthesise results. | Yes |
| **RESULTS** | | |  |
| Included studies | 7 | Give the total number of included studies and participants and summarise relevant characteristics of studies. | Yes |
| Synthesis of results | 8 | Present results for main outcomes, preferably indicating the number of included studies and participants for each. If meta-analysis was done, report the summary estimate and confidence/credible interval. If comparing groups, indicate the direction of the effect (i.e. which group is favoured). | Yes |
| **DISCUSSION** | | |  |
| Limitations of evidence | 9 | Provide a brief summary of the limitations of the evidence included in the review (e.g. study risk of bias, inconsistency and imprecision). | Yes |
| Interpretation | 10 | Provide a general interpretation of the results and important implications. | Yes |
| **OTHER** | | |  |
| Funding | 11 | Specify the primary source of funding for the review. | Yes |
| Registration | 12 | Provide the register name and registration number. | Yes |

From:  Page MJ, McKenzie JE, Bossuyt PM, Boutron I, Hoffmann TC, Mulrow CD, et al. The PRISMA 2020 statement: an updated guideline for reporting systematic reviews. BMJ 2021;372:n71. doi: 10.1136/bmj.n71. This work is licensed under CC BY 4.0. To view a copy of this license, visit [https://creativecommons.org/licenses/by/4.0/](https://creativecommons.org/licenses/by/4.0/ )

### 2. GATHER Statement

#### Table S2. GATHER (Guidelines for Accurate and Transparent Health Estimates Reporting) statement checklist

| **Item #** | **Checklist item** | **Reported on page #** |
| --- | --- | --- |
| **Objectives and funding** | | |
| **1** | Define the indicator(s), populations (including age, sex, and geographic entities), and  time period(s) for which estimates were made. | P 4 |
| **2** | List the funding sources for the work. | P 2 |
| **Data Inputs** | | |
| *For all data inputs from multiple sources that are synthesized as part of the study:* | | |
| **3** | Describe how the data were identified and how the data were accessed. | P 5,6 |
| **4** | Specify the inclusion and exclusion criteria. Identify all ad‐hoc exclusions. | P 5,6 and Appendix |
| **5** | Provide information on all included data sources and their main characteristics. For each data source used, report reference information or contact name/institution, population represented, data collection method, year(s) of data collection, sex and age range,  diagnostic criteria or measurement method, and sample size, as relevant. | Appendix Table S3, Supplementary file |
| **6** | Identify and describe any categories of input data that have potentially important biases  (e.g., based on characteristics listed in item 5). | Appendix P 13 |
| *For data inputs that contribute to the analysis but were not synthesized as part of the study:* | | |
| **7** | Describe and give sources for any other data inputs. | N/A |
| *For all data inputs:* | | |
| **8** | Provide all data inputs in a file format from which data can be efficiently extracted (e.g., a spreadsheet rather than a PDF), including all relevant meta‐data listed in item 5. For any data inputs that cannot be shared because of ethical or legal reasons, such as third‐party ownership, provide a contact name or the name of the institution that retains the right to  the data. | Appendix Table S3, Supplementary file |
| **Data analysis** | | |
| **9** | Provide a conceptual overview of the data analysis method. A diagram may be helpful. | P 6 |
| **10** | Provide a detailed description of all steps of the analysis, including mathematical formulae. This description should cover, as relevant, data cleaning, data pre‐processing, data adjustments and weighting of data sources, and mathematical or statistical  model(s). | P 6,7 |
| **11** | Describe how candidate models were evaluated and how the final model(s) were  selected. | P 7 |
| **12** | Provide the results of an evaluation of model performance, if done, as well as the results  of any relevant sensitivity analysis. | P 7 |
| **13** | Describe methods for calculating uncertainty of the estimates. State which sources of  uncertainty were, and were not, accounted for in the uncertainty analysis. | P 7 |
| **14** | State how analytic or statistical source code used to generate estimates can be accessed. | The analytic/statistical code used to generate the estimates is available from the corresponding author upon reasonable request. |
| **Results and Discussion** | | |
| **15** | Provide published estimates in a file format from which data can be efficiently extracted. | Supplementary file  Appendix Table S3 |
| **16** | Report a quantitative measure of the uncertainty of the estimates (e.g. uncertainty  intervals). | Table 1-2 & Appendix Table S6 |
| **17** | Interpret results in light of existing evidence. If updating a previous set of estimates,  describe the reasons for changes in estimates. | P 8 |
| **18** | Discuss limitations of the estimates. Include a discussion of any modelling assumptions or  data limitations that affect interpretation of the estimates. | P 9 |

### 3. Stage 1- National-level data source

#### Table S3. Countries with available national or administrative data on induction of labour and corresponding data sources

| **Country** | **Data Source** | **URL** | **Year of data reported** |
| --- | --- | --- | --- |
| Austria | Birth Register Austria Annual Reports | <https://www.iet.at/page.cfm?vpath=register/geburtenregister> | 2010-2021 |
| Belgium | Performance of the Belgium Health System KCE 2019 report | <https://www.healthybelgium.be/metadata/hspa/mn4.pdf> | 2010-2015 |
| Cyprus | Perinatal Health report, Ministry of Health | [https://www.gov.cy/moh/en/documents/health-monitoring-unit-statistics/perinatal-health/publications/](https://www.gov.cy/moh/en/documents/health-monitoring-unit-statistics/perinatal-health/publications/ ) | 2010-2016 |
| Estonia | Health Statistics and Health Research Database | <https://statistika.tai.ee/pxweb/en/Andmebaas/Andmebaas__01Rahvastik__02Synnid/SR64.px/> (2010-2019)  <https://statistika.tai.ee/pxweb/en/Andmebaas/Andmebaas__01Rahvastik__02Synnid/SR641.px/> (2020-2022)  Contact:  | 2010-2022 |
| Finland | Finish Institute for Health and Welfare | [https://sampo.thl.fi/pivot/prod/en/synre/toimenpiteet/summary_timebar?sairaala_0=10822&aiemmatsynnytykset_0=10768&sikioisyys_0=10837&ika_0=10452&raskausviikot_0=10711&bmi_0=73019&mittarit_0=10870](https://sampo.thl.fi/pivot/prod/en/synre/toimenpiteet/summary_timebar?sairaala_0=10822&aiemmatsynnytykset_0=10768&sikioisyys_0=10837&ika_0=10452&raskausviikot_0=10711&bmi_0=73019&mittarit_0=10870 ) | 2010-2021 |
| Germany | Federal evaluation for the data collection/Obstetric quality indicators and key figures;  Temporal trends in fetal mortality at and beyond term and induction of labor in Germany 2005–2012: data from German routine perinatal monitoring | <https://www.ncbi.nlm.nih.gov/pmc/articles/PMC4709369/pdf/404_2015_Article_3795.pdf> (2010-2012)  <https://iqtig.org/qs-verfahren/peri/> (2016-2019) | 2010-2012,  2016-2019 |
| Iceland | Iceland Birth Registrations report | <https://island.is/en/faedingar-tolur/arsskyrslur-faedingaskraningar>  Contact:  | 2010-2020 |
| Italy | Certificate of Birth Assistance CeDAP | [https://www.salute.gov.it/new/it/scheda-statistica/certificato-di-assistenza-al-parto-cedap-analisi-dellevento-nascita/](https://www.salute.gov.it/new/it/scheda-statistica/certificato-di-assistenza-al-parto-cedap-analisi-dellevento-nascita/ ) | 2010-2022 |
| Lithunia | Statistics Lithuania (Official Statistics Portal) | [https://www.hi.lt/sveikatos-statistikos-leidiniai/#--gimimu-medicininiai-duomenys](https://www.hi.lt/sveikatos-statistikos-leidiniai/%23--gimimu-medicininiai-duomenys ) | 2010-2021 |
| Luxembourg | Perinatal Health Surveillance in Luxembourg | [https://santesecu.public.lu/fr/espace-professionnel/informations-donnees/perinat.html](https://santesecu.public.lu/fr/espace-professionnel/informations-donnees/perinat.html ) | 2010-2019 |
| Malta | National Obstetric Information System Annual Report | [https://dhir.gov.mt/en/resources/birth-reports/](https://dhir.gov.mt/en/resources/birth-reports/ ) | 2010, 2013-2019, 2021 |
| Netherlands | Netherlands Perinatal Registry (PRN) | <https://www.europeristat.com/wp-content/uploads/2013/05/EPHR2010_w_disclaimer.pdf> (page 230 for 2010)  [Peristat.nl](https://www.peristat.nl/) (2012-2021) | 2010, 2012-2021 |
| Norway | Norwegian Institute of Health, Medical Birth Register | [https://statistikk.fhi.no/mfr/Kg8Qeund8-FnrmTJlQhiN0Hfz6KiPTGOefdLvHp3aJA](https://statistikk.fhi.no/mfr/Kg8Qeund8-FnrmTJlQhiN0Hfz6KiPTGOefdLvHp3aJA ) | 2010-2022 |
| Spain | Perinatal care in Spain: analysis of physical and human resources, activity and quality of hospital services 2010-2018. Ministry of Health Spain (Figure 22) | <https://www.sanidad.gob.es/estadEstudios/estadisticas/docs/Informe_Atencion_Perinatal_2010-2018.pdf> | 2010-2018 |
| Sweden | Statistics on pregnancies, childbirths and newborns, sicialstyrelsen | <https://www.socialstyrelsen.se/statistik-och-data/statistik/alla-statistikamnen/graviditeter-forlossningar-och-nyfodda/> | 2010-2021 |
| England | NHS Maternity Statistics, Chapter Deliveries over time; Method of onset | <https://digital.nhs.uk/data-and-information/publications/statistical/nhs-maternity-statistics> | 2010-2022 |
| USA | Centres for Disease Control and Prevention, CDC WONDER | <https://www.cdc.gov/nchs/data_access/VitalStatsOnline.htm#Births> (2010-2015)  <http://wonder.cdc.gov/natality-expanded-current.html> (2016-2022)  Contact:  | 2010-2022 |
| Australia | Australian mothers and babies report, Australia Institute of Health and Welfare (AIHW) | [https://www.aihw.gov.au/reports/mothers-babies/australias-mothers-babies/contents/labour-and-birth/onset-of-labour](https://www.aihw.gov.au/reports/mothers-babies/australias-mothers-babies/contents/labour-and-birth/onset-of-labour ) | 2010-2021 |
| New Zealand | Ministry of Health New Zealand  Labour and Birth: Selected interventions by year and district | <https://tewhatuora.shinyapps.io/report-on-maternity-web-tool/> | 2010-2021 |

### 4. Stage 2- Published literature search

#### 4.1 Search strategy

**Database: Ovid MEDLINE(R) <1946 to June 13, 2024>**  
**Search Strategy:**  
**1**  labor, induced/ or amniotomy/ (10414)  
**2**  (induc* adj2 labo?r).mp. (14145)  
**3**  amniotom*.mp. (774)  
**4**  artificial rupture of membranes.mp. (150)  
**5**  deliberate rupture of membranes.mp. (1)  
**6**  (labo?r acceleration or "acceleration of labo?r" or parturition induction or "induction of parturition" or stimulated labo?r or "stimulation of labo?r").mp. (441)  
**7**  Delivery, Obstetric/ (33886)  
**8**  ((mode adj2 deliver*) or assisted deliver* or (type* adj2 delivery) or vaginal deliver* or c?esarian or labo?r complication*).mp. (43674)  
**9**  7 and 8 (10325)  
**10**  1 or 2 or 3 or 4 or 5 or 6 or 9 (23988)  
**11**  Epidemiology/ (12600)  
**12**  data collection/ or datasets as topic/ or records/ or registries/ or incidence/ or prevalence/ or epidemiological monitoring/ or sentinel surveillance/ or statistics as topic/ (923180)  
**13**  (data set* or dataset* or records or registries or register* or registry or incidence or prevalence or epidemiolog* or surveillance or statistics).mp. (4785378)  
**14**  (nationwide or nation wide or countrywide or country-wide or world wide or worldwide or population level or population wide or hospital based).mp. (400445)  
**15**  ((global or world or region* or country or nation*) adj3 (estimate* or rate* or level* or data* or survey*)).mp. (261340)  
**16**  ((population-based, community-based or facility based) adj3 (data or estimate* or rate* or level* or survey*)).mp. (363)  
**17**  11 or 12 or 13 or 14 or 15 or 16 (5117445)  
**18**  10 and 17 (9934)  
**19**  Labor, Induced/sn, td [Statistics & Numerical Data, Trends] (872)  
**20**  amniotomy/sn, td (9)  
**21**  (induc* adj2 labo?r adj3 (rate* or number*)).mp. (488)  
**22**  18 or 19 or 20 or 21 (10106)  
**23**  (Austria* or Belgium* or Brussel* or Flander* or Wallonia* or Cyprus* or Estonia* or Finland* or German* or Iceland* or Italy* or Lithuania* or Luxembourg* or Malta* or Netherland* or Norway* or Spain* or Sweden* or England* or Scotland* or Wales* or United State* or USA or North America or Australia* or New Zealand*).mp. (2470079)  
**24**  22 not 23 (8044)  
**25**  limit 24 to yr="2010 -Current" (4741)  
**26**  exp animals/ not humans.sh. (5230954)  
**27**  25 not 26 (4713) 

**Database: APA PsycInfo <1806 to June Week 1 2024>**  
**Search Strategy:**  
**1**  (artificial rupture of membranes or deliberate rupture of membranes).mp. (3)  
**2**  (induc* adj2 labo?r).mp. (285)  
**3**  amniotom*.mp. (11)  
**4**  (labo?r acceleration or "acceleration of labo?r" or parturition induction or "induction of parturition" or stimulated labo?r or "stimulation of labo?r").mp. (7)  
**5**  ((mode adj2 deliver*) or assisted deliver* or (type* adj2 delivery) or vaginal deliver* or c?esarian or labo?r complication*).mp. (2628)  
**6**  obstetrics/ (1636)  
**7**  5 and 6 (132)  
**8**  1 or 2 or 3 or 4 or 7 (420)  
**9**  epidemiology/ (57709)  
**10**  register/ or incidence/ or prevalence/ or monitoring/ or population statistics/ or statistics/ or vital statistics as topic/ (21964)  
**11**  (data set* or dataset* or records or registries or register* or registry or incidence or prevalence or epidemiolog* or surveillance or statistics).mp. (523052)  
**12**  (nationwide or nation wide or countrywide or country-wide or world wide or worldwide or population level or population wide or hospital based).mp. (57208)  
**13**  ((global or world or region* or country or nation*) adj3 (estimate* or rate* or level* or data* or survey*)).mp. (108481)  
**14**  ((population-based or community-based or facility based) adj3 (data or estimate* or rate* or level* or survey*)).mp. (5881)  
**15**  9 or 10 or 11 or 12 or 13 or 14 (647394)  
**16**  8 and 15 (156)  
**17**  (induc* adj2 labo?r adj3 (rate* or number*)).mp. (23)  
**18**  ((acceleration or stimulation) adj2 labo?r adj3 (rate* or number*)).mp. (0)  
**19**  16 or 17 or 18 (168)  
**20**  (Austria* or Belgium* or Brussel* or Flander* or Wallonia* or Cyprus* or Estonia* or Finland* or German* or Iceland* or Italy* or Lithuania* or Luxembourg* or Malta* or Netherland* or Norway* or Spain* or Sweden* or England* or Scotland* or Wales* or United State* or USA or North America or Australia* or New Zealand*).mp. (543026)  
**21**  19 not 20 (112)  
**22**  limit 21 to yr="2010 -Current" (76)  
**23**  limit 22 to human (68) 

**Database: Embase Classic+Embase <1947 to 2024 June 13>**  
**Search Strategy:**  
**1**  labor induction/ or amniotomy/ (18931)  
**2**  (artificial rupture of membranes or deliberate rupture of membranes).mp. (288)  
**3**  (induc* adj2 labo?r).mp. (25125)  
**4**  amniotom*.mp. (1832)  
**5**  (labo?r acceleration or "acceleration of labo?r" or parturition induction or "induction of parturition" or stimulated labo?r or "stimulation of labo?r").mp. (554)  
**6**  1 or 2 or 3 or 4 or 5 (26233)  
**7**  ((mode adj2 deliver*) or assisted deliver* or (type* adj2 delivery) or vaginal deliver* or c?esarian or labo?r complication*).mp. (85263)  
**8**  obstetric delivery/ (24139)  
**9**  7 and 8 (4305)  
**10**  6 or 9 (30075)  
**11**  epidemiology/ (286699)  
**12**  register/ or incidence/ or prevalence/ or monitoring/ or population statistics/ or statistics/ or vital statistics as topic/ (2134065)  
**13**  (data set* or dataset* or records or registries or register* or registry or incidence or prevalence or epidemiolog* or surveillance or statistics).mp. (6146113)  
**14**  (nationwide or nation wide or countrywide or country-wide or world wide or worldwide or population level or population wide or hospital based).mp. (674948)  
**15**  ((global or world or region* or country or nation*) adj3 (estimate* or rate* or level* or data* or survey*)).mp. (437520)  
**16**  ((population-based or community-based or facility based) adj3 (data or estimate* or rate* or level* or survey*)).mp. (39259)  
**17**  11 or 12 or 13 or 14 or 15 or 16 (6878565)  
**18**  10 and 17 (8746)  
**19**  (induc* adj2 labo?r adj3 (rate* or number*)).mp. (876)  
**20**  ((acceleration or stimulation) adj2 labo?r adj3 (rate* or number*)).mp. (6)  
**21**  18 or 19 or 20 (9245)  
**22**  (Austria* or Belgium* or Brussel* or Flander* or Wallonia* or Cyprus* or Estonia* or Finland* or German* or Iceland* or Italy* or Lithuania* or Luxembourg* or Malta* or Netherland* or Norway* or Spain* or Sweden* or England* or Scotland* or Wales* or United State* or USA or North America or Australia* or New Zealand*).mp. (4594689)  
**23**  21 not 22 (7444)  
**24**  (exp animal/ or nonhuman/ or exp invertebrate/ or animal.hw.) not exp human/ (8279673)  
**25**  23 not 24 (7331)  
**26**  limit 25 to yr="2010 -Current" (5432) 

**Database: SCOPUS 13th June (5624)**

((((TITLE-ABS-KEY(("data set*" or dataset* or records or registries or register* or registry or incidence or prevalence or epidemiolog* or surveillance or statistics)) OR TITLE-ABS-KEY((nationwide or "nation wide" or countrywide or "country-wide" or "world wide" or worldwide or "population level" or "population wide" or "hospital based")) OR TITLE-ABS-KEY(((global or world or region* or country or nation*) W/2 (estimate* or rate* or level* or data* or survey*))) OR TITLE-ABS-KEY((("population-based" or "community-based" or "facility based") W/2 (data or estimate* or rate* or level* or survey*))))) AND (((TITLE-ABS-KEY((induc* W/1 labo?r)) OR TITLE-ABS-KEY((amniotom* or "artificial rupture of membranes" or "deliberate rupture of membranes")) OR TITLE-ABS-KEY(("labo?r acceleration" or "acceleration of labo?r" or "parturition induction" or "induction of parturition" or "stimulated labo?r" or "stimulation of labo?r")))) OR ((TITLE-ABS-KEY(((mode W/1 deliver*) or "assisted deliver*" or (type* W/1 delivery) or "vaginal deliver*" or c?esarian or "labo?r complication*")) AND TITLE-ABS-KEY("obstetric delivery"))))) OR ((TITLE-ABS-KEY((induc* W/1 labo?r W/2 (rate* or number*))) OR TITLE-ABS-KEY(((acceleration or stimulation) W/1 labo?r W/2 (rate* or number*)))))) AND NOT (TITLE-ABS-KEY((Austria* or Belgium* or Brussel* or Flander* or Wallonia* or Cyprus* or Denmark* or Estonia* or Finland* or France* or French* or German* or Iceland* or Italy* or Lithuania* or Luxembourg* or Malta* or Netherland* or Norway* or Sweden* or England* or Scotland* or Wales* or "United State*" or USA or North America or Australia* or "New Zealand*"))) AND ( LIMIT-TO ( PUBYEAR,2010) OR LIMIT-TO ( PUBYEAR,2011) OR LIMIT-TO ( PUBYEAR,2012) OR LIMIT-TO ( PUBYEAR,2013) OR LIMIT-TO ( PUBYEAR,2014) OR LIMIT-TO ( PUBYEAR,2015) OR LIMIT-TO ( PUBYEAR,2016) OR LIMIT-TO ( PUBYEAR,2017) OR LIMIT-TO ( PUBYEAR,2018) OR LIMIT-TO ( PUBYEAR,2019) OR LIMIT-TO ( PUBYEAR,2020) OR LIMIT-TO ( PUBYEAR,2021) OR LIMIT-TO ( PUBYEAR,2022) OR LIMIT-TO ( PUBYEAR,2023) OR LIMIT-TO ( PUBYEAR,2024) )

**Database: CINAHL Search Strategy (EBSCOhost)> June 15, 2024**

S1: (MH "labor, induced") OR (MH amniotomy)

S2: (induc* N2 labo#r)

S3: amniotom*

S4: "artificial rupture of membranes"

S5: "deliberate rupture of membranes"

S6: (MH "Delivery, Obstetric")

S7: ((mode N2 deliver*) OR "assisted deliver*" OR (type* N2 delivery) OR "vaginal deliver*" OR c#esarian OR "labo#r complication*")

S8: S6 AND S7

S9: S1 OR S2 OR S3 OR S4 OR S5 OR S8

S10: (MH Epidemiology)

S11: (MH "data collection") OR (MH "datasets as topic") OR (MH records) OR (MH registries") OR (MH incidence) OR (MH prevalence)

OR (MH "epidemiological monitoring") OR (MH "sentinel surveillance") OR (MH "statistics as topic")

S12: ("data set*" OR dataset* OR records OR registries OR register* OR registry OR incidence OR prevalence OR epidemiolog* OR surveillance

OR statistics) OR (nationwide OR "nationwide" OR countrywide OR "worldwide" OR worldwide OR "population level" OR "population wide"

OR "hospital based")

S13: ((global OR world OR region* OR country OR nation*) N3 (estimate* OR rate* OR level* OR data* OR survey*))

S14: (("population-based" OR "community-based" OR "facility based") N3 (data OR estimate* OR rate* OR level* OR survey*))

S15: S10 OR S11 OR S12 OR S13 OR S14

S16: S9 AND S15

S17: (Austria* OR Belgium* OR Brussel* OR Flander* OR Wallonia* OR Cyprus* OR Estonia* OR Finland* OR German* OR Iceland*

OR Italy* OR Lithuania* OR Luxembourg* OR Malta* OR Netherland* OR Norway* OR Spain* OR Spanish* OR Sweden* OR England*

OR Scotland* OR Wales* OR "United State*" OR USA OR North America* OR Australia* OR "New Zealand*")

S18: S16 NOT S17

S19: S16 NOT S17

#### 4.2 Definition of high-risk pregnancy

Any medical or obstetric conditions during pregnancy that may pose potential harm or hazard to the mother or foetus are considered as high-risk pregnancy.^1^ This systematic review will follow the categories outlined by National Institute of Health^2^  that may result in high-risk pregnancies. Studies conducted on pregnant women that only include the following conditions will be excluded:

1. Existing health conditions: Hypertensive disorders (chronic/pre-existing hypertension), polycystic ovarian syndrome, diabetes, renal disease, autoimmune disease, thyroid disease, infertility, obesity, HIV/AIDS
2. Age: Adolescent or first-time pregnancy after 35 years of age
3. Lifestyle factors: Alcohol use, tobacco use, illicit drug use (during pregnancy)
4. Conditions of pregnancy: Multiple gestation, Gestational diabetes, Preeclampsia and eclampsia, gestational hypertension.

Additionally, pregnant women with conditions such as thrombophilia (congenital and acquired), fetal growth restriction, antepartum haemorrhage and mental disorder (anxiety, depression, bipolar disorder, PPD) will not be considered. These conditions are identified as high-risk by a 2017 Cochrane review.^3^

#### 4.3 Methodological approach to selecting studies in stage 2

Stage 2 identified a total of N=176 eligible studies reporting data on labour induction for our period of interest (2010 to 2023).

Studies reporting induction data for a single year were included for that year.

For studies reporting induction data across multi-year periods:

- If year-specific data were reported, separate datapoints corresponding to each year were extracted.
- If year-specific data were not provided, and only a single overall induction rate was presented for the entire multi-year period, we assigned the reported rate to the mid-year of that period. For example, an overall rate of 20% reported for 2014–2019 was treated as the 2016 rate.
- We identified studies that were published after 2010, with multi-year data collection periods that started prior to 2010. In this situation, we included only those studies where at least part of the study period overlapped with 2010 or later. For these studies:
  - If year-specific data were reported for the years 2010 onwards, separate datapoints corresponding to each eligible year were extracted.
  - If year-specific data were not provided (i.e. only a single overall induction rate was presented for the entire multi-year period):
    - If we had identified another study that reported data for the same year/s for that country, the study was excluded.
    - If no other studies had reported data for that country for the post-2010 data collection period, the study was retained and the multi-year rate applied.
- We identified many studies reporting data for the same country in the same year (described here as country-year), which in some instances were two studies using the same dataset. This carries a risk of data duplication. We aimed to minimise these risks, while ensuring the most comprehensive and representative data were included in the dataset for analysis. We therefore developed a stepwise process to identify a single, best datapoint per country-year to include in the database for analysis:

1. For a given country-year, if only one eligible study was identified, it was included.
2. If multiple studies for the same country-year were identified, we applied a hierarchical selection to identify the most useful datapoints to be included for analysis.
3. **Step 1: Data source quality.** Studies were categorised in the following way:
   - Category A: National or state administrative data, national birth or perinatal registries/surveys
   - Category B: Data from multiple facilities or administrative units within a country
   - Category C: Single-centre data

We selected the study with the highest available category (A > B > C). If only one study remained after Step 1, then that study was selected. If multiple studies remained (for example, two studies in Category C) we moved to Step 2.

4. **Step 2. Time period.** If one study reported specific, annual data (i.e. country-year), we selected this over another study that reported a proportion over multiple years (for example an assumed average induction rate of 20% for 2013-2016). If multiple studies reported proportions over multiple years, we selected the study with the narrowest time period.
5. **Step 3. Sample size.** If multiple studies remained after Steps 1 and 2 (for example, studies within the same data source category, reporting induction data for the same year), we selected the study with the largest sample size.

### 5. Countries included in 2019 estimate and trend analysis

#### 5.1 Table S4. List of countries included in the analysis

| **Country** | **Income Level** | **WHO Region** | **Inclusion in the analysis** |
| --- | --- | --- | --- |
| Austria | High income | European Region | ☒ 2019 Estimate  ☒ Trend Analysis |
| Australia | High income | Western Pacific Region | ☒ 2019 Estimate  ☒ Trend Analysis |
| Belgium | High income | European Region | ☐ 2019 Estimate  ☒ Trend Analysis |
| Brazil | Upper middle income | Region of America | ☒ 2019 Estimate  ☒ Trend Analysis |
| Canada | High income | Region of America | ☒ 2019 Estimate  ☒ Trend Analysis |
| China | Upper middle  income | Western Pacific Region | ☒ 2019 Estimate  ☒ Trend Analysis |
| Croatia | High income | European Region | ☒ 2019 Estimate  ☒ Trend Analysis |
| Cyprus | High income | European Region | ☐ 2019 Estimate  ☒ Trend Analysis |
| Czechia (Czech Republic) | High income | European Region | ☒ 2019 Estimate  ☒ Trend Analysis |
| Denmark | High income | European Region | ☒ 2019 Estimate  ☒ Trend Analysis |
| England | High income | European Region | ☒ 2019 Estimate  ☒ Trend Analysis |
| Estonia | High income | European Region | ☒ 2019 Estimate  ☒ Trend Analysis |
| Finland | High income | European Region | ☒ 2019 Estimate  ☒ Trend Analysis |
| France | High income | European Region | ☒ 2019 Estimate  ☒ Trend Analysis |
| Germany | High income | European Region | ☒ 2019 Estimate  ☒ Trend Analysis |
| Hungary | High income | European Region | ☒ 2019 Estimate  ☒ Trend Analysis |
| Iceland | High income | European Region | ☒ 2019 Estimate  ☒ Trend Analysis |
| India | Lower middle income | South-East Asian Region | ☒ 2019 Estimate  ☒ Trend Analysis |
| Iran | Lower middle income | Eastern Mediterranean Region | ☒ 2019 Estimate  ☒ Trend Analysis |
| Ireland | High income | European Region | ☒ 2019 Estimate  ☒ Trend Analysis |
| Israel | High income | European Region | ☒ 2019 Estimate  ☒ Trend Analysis |
| Italy | High income | European Region | ☒ 2019 Estimate  ☒ Trend Analysis |
| Japan | High income | Western Pacific Region | ☒ 2019 Estimate  ☒ Trend Analysis |
| Latvia | High income | European Region | ☒ 2019 Estimate  ☒ Trend Analysis |
| Lithuania | High income | European Region | ☒ 2019 Estimate  ☒ Trend Analysis |
| Luxembourg | High income | European Region | ☒ 2019 Estimate  ☒ Trend Analysis |
| Malaysia | Upper middle  income | Western Pacific Region | ☒ 2019 Estimate  ☒ Trend Analysis |
| Malta | High income | European Region | ☒ 2019 Estimate  ☒ Trend Analysis |
| Netherlands | High income | European Region | ☒ 2019 Estimate  ☒ Trend Analysis |
| New Zealand | High income | Western Pacific Region | ☒ 2019 Estimate  ☒ Trend Analysis |
| Norway | High income | European Region | ☒ 2019 Estimate  ☒ Trend Analysis |
| Poland | High income | European Region | ☒ 2019 Estimate  ☒ Trend Analysis |
| Portugal | High income | European Region | ☒ 2019 Estimate  ☒ Trend Analysis |
| Qatar | High income | Eastern Mediterranean Region | ☒ 2019 Estimate  ☒ Trend Analysis |
| Saudi Arabia | High income | Eastern Mediterranean Region | ☒ 2019 Estimate  ☒ Trend Analysis |
| Slovenia | High income | European Region | ☒ 2019 Estimate  ☒ Trend Analysis |
| Spain | High income | European Region | ☐ 2019 Estimate  ☒ Trend Analysis |
| Sweden | High income | European Region | ☒ 2019 Estimate  ☒ Trend Analysis |
| Switzerland | High income | European Region | ☒ 2019 Estimate  ☒ Trend Analysis |
| Turkey | Upper middle  income | European Region | ☒ 2019 Estimate  ☒ Trend Analysis |
| United Arab Emirates | High income | Eastern Mediterranean Region | ☒ 2019 Estimate  ☒ Trend Analysis |
| United States | High income | Region of America | ☒ 2019 Estimate  ☒ Trend Analysis |
| Uruguay | High income | Region of America | ☒ 2019 Estimate  ☒ Trend Analysis |
